# MedAgent: A Retrieval-Augmented Clinical Decision Support Agent with Verifiable Evidence Grounding for Evidence-Based Medicine

**DOI:** 10.64898/2026.06.15.26355735

**Authors:** Fuqiang Wang, Zhicai Guo, Zhikang Ye

**Author notes:** Equal contribution (Fuqiang Wang, Zhicai Guo).

## Abstract

Evidence-based medicine demands clinical answers that are not only fluent and medically plausible, but also anchored in traceable evidence, tailored to patient-specific clinical questions, sensitive to the hierarchy of evidence, and respectful of clinical safety boundaries. While general-purpose large language models (LLMs) exhibit strong medical language generation ability, they tend to lean on parametric memory, underuse retrieved evidence, hallucinate citations, conflate evidence levels, and draw conclusions that are not fully supported by the underlying literature. Such limitations pose particular risks in clinical decision support, where answer reliability, evidence traceability, and reasoning consistency are paramount.

To address these issues, we present MedAgent, an evidence-based medical agent trained through an end-to-end pipeline that integrates supervised fine-tuning (SFT) cold start, reward modeling, and Group Relative Policy Optimization (GRPO). The agent is designed to execute a structured workflow encompassing clinical question understanding, PICO extraction, evidence retrieval, evidence stratification, citation-grounded answer generation, and quality evaluation. Specifically, a Qwen2.5-14B-Instruct backbone is first cold-started on 200 human-verified agent trajectories, equipping it with tool invocation, PICO parsing, structured response generation, and citation faithfulness. Next, a Qwen2.5-7B reward model is trained on 2,099 pairwise preference samples to provide semantic-level quality signals for evidence-based responses. Finally, GRPO reinforcement learning is conducted in a retrieval-augmented agent environment, where every rollout involves real evidence retrieval and is scored jointly by rule-based rewards and reward-model signals.

To avoid over-reliance on training rewards, we further construct an independent evidence-based medical evaluation benchmark, MedTrustBench, which contains 200 clinical questions spanning 10 specialties and four difficulty levels. Each question is annotated with standardized PICO elements and rubric-based scoring criteria. The benchmark includes 1,187 rubrics across seven dimensions: question relevance, evidence hierarchy, evidence quality and timeliness, evidence-answer consistency, completeness and depth, logical rigor, and medical terminology. Under an identical RAG pipeline, retrieval tool, retrieval configuration, and evaluation protocol, MedAgentv17 attains 78.6 points, outperforming GPT-4.1 (75.3) and approaching GPT-5.4 (80.3). These results show that a 14B domain-aligned model can surpass strong general-purpose baselines on specialized evidence-based medical reasoning, while delivering practical advantages in cost, privacy, controllability, and hospital-oriented private deployment. The model and associated datasets are publicly released at https://www.modelscope.cn/profile/InfoxmedModel.

## I. Introduction

Large language models (LLMs) [1]–[4] have delivered remarkable progress in medical question answering, clinical text summarization, and biomedical knowledge generation [5]–[7]. Yet clinical evidence-based decision support differs substantially from general medical dialogue. In real clinical settings, a useful answer must do more than offer a plausible recommendation: it must also identify which evidence underpins the recommendation, characterize the level of that evidence, assess its reliability, judge whether the conclusion generalizes to the specific patient population, and acknowledge any remaining uncertainty or conflicting evidence [8], [9].

Evidence-based medicine centers on the integration of clinical expertise, patient-specific conditions, and the best available research evidence [8]. A widely adopted approach for formulating clinical questions is the PICO framework, which decomposes a question into Population, Intervention, Comparison, and Outcome [10]. Beyond understanding the clinical question, an evidence-based medical assistant should retrieve relevant literature from multiple evidence sources, including clinical guidelines, systematic reviews, meta-analyses, and randomized controlled trials (RCTs) [11], [12]. It should then organize the evidence by hierarchy [9], [12] and produce a traceable response with faithful citations.

Directly applying general-purpose LLMs to this task proves inadequate [13], [14]. Although such models can generate fluent medical explanations [5], they exhibit several recurring limitations in strict evidence-based settings. First, they tend to fall back on parametric memory and answer without invoking retrieval tools [15], [16]. Second, even when retrieval results are provided, they may underuse the retrieved evidence or produce conclusions inconsistent with the retrieved documents [17], [18]. Third, they may fabricate citation identifiers, clinical trials, effect sizes, or guideline recommendations [13], [14], [19]. Fourth, they may blur evidence levels, treating expert opinion, guidelines, systematic reviews, and RCTs as if they carried equal evidential weight [9]. Finally, they may issue overconfident recommendations when evidence is incomplete or conflicting.

In ordinary question answering these issues mainly degrade answer quality, but in medical scenarios they directly compromise trustworthiness, interpretability, and clinical safety [6], [20]. Accordingly, the goal of this work is not simply to teach a model to “speak medical language better”, but to train it to behave as a genuine evidence-based medical agent [21], [22]. The agent should follow a stable workflow:

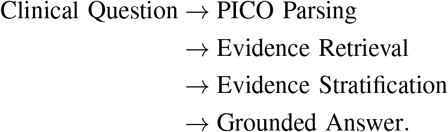

To this end, we propose MedAgent, a domain-specialized medical agent trained through a multi-stage alignment pipeline that comprises three core stages. First, supervised fine-tuning (SFT) [23], [24] serves as a cold-start stage that instills agent-level behaviors, including tool invocation, PICO extraction, structured answer generation, and citation faithfulness. Second, a reward model is trained on pairwise preference data to distinguish high-quality evidence-based answers from weaker ones [23], [25], [26]. Third, GRPO reinforcement learning [27], [28] is conducted in an online retrieval-augmented environment, in which each rollout requires the model to call retrieval tools, ingest the returned evidence, and produce structured answers.

Nevertheless, the convergence of training rewards alone is no guarantee of clinical reliability. A high reward value merely signals that the policy has learned to produce outputs favored by the reward function or reward model; it does not directly answer the questions that matter most in clinical practice: Are the cited data accurate? Are any trials or statistics hallucinated? Does the model behave safely under evidence conflict? Does it organize evidence according to the evidence-based medical hierarchy? Does it refrain from overconfident recommendations when evidence is insufficient? Answering these questions calls for an independent, dimension-specific evaluation benchmark [20], [29].

To this end, we further construct MedTrustBench, an evidence-based medical evaluation benchmark that is independent of the training process. It comprises 200 clinical decision questions, covers 10 medical specialties, and adopts rubric-based evaluation in place of fixed gold answers, thereby complementing existing medical QA benchmarks such as MedQA [30], PubMedQA [31], and MMLU [32]. Each question is annotated with PICO elements, difficulty level, specialty label, source literature, and multiple verifiable rubrics. The rubrics are formulated as Yes/No-style criteria with positive and negative weights, supporting both automated scoring and fine-grained diagnostic analysis.

The main contributions of this paper are summarized as follows:

- We propose MedAgent, an evidence-based medical agent trained through SFT cold start, reward modeling, and GRPO reinforcement learning.
- We curate a high-quality SFT cold-start dataset of 200 human-verified agent trajectories that enable stable tool calling, PICO extraction, structured evidence-based answering, and disciplined citation behavior.
- We train a Qwen2.5-7B reward model on 2,099 pairwise preference samples to deliver semantic-level quality signals for medical evidence-based response optimization.
- We design a hybrid reward function that couples rule-based constraints with reward-model scores, explicitly promoting tool use, PICO completeness, citation faithfulness, evidence hierarchy, and clinical safety.
- We build MedTrustBench, an independent rubric-based benchmark featuring 200 clinical questions, 10 specialties, four difficulty levels, and 1,187 rubrics across seven evaluation dimensions.
- Experiments demonstrate that MedAgentv17 reaches 78.6 points under the same RAG pipeline, exceeding GPT-4.1 by 3.3 points and narrowing the gap to GPT-5.4 to just 1.7 points.

## II. Related Work

### A. Medical Large Language Models

A rapidly growing body of work investigates how to adapt LLMs to medicine. Med-PaLM and Med-PaLM 2 [5], [6] show that PaLM-scale models with medical instruction tuning can approach expert-level performance on USMLE-style questions, while GPT-4 has been reported to reach state-of-the-art results on a broad range of medical benchmarks [2], [7]. Open medical LLMs include MEDITRON [33], which scales medical pretraining to 70B parameters; PMC-LLaMA [34], which combines medical corpus pretraining with instruction tuning; ChatDoctor [35], Clinical Camel [36], and ClinicalGPT [37]; as well as HuatuoGPT and HuatuoGPT-o1 [38], [39] targeting Chinese medical scenarios. Whereas these models primarily optimize for medical knowledge or dialogue ability, MedAgent emphasizes the *evidence-based behavior* of a medical agent: tool calling, PICO parsing, citation-faithful answering, and evidence stratification.

### B. Retrieval-Augmented Generation in Medicine

Retrieval-augmented generation (RAG) [15], [16] couples parametric LLMs with non-parametric retrieval and has become a standard recipe for knowledge-intensive tasks. Dense retrievers such as DPR [40] and classical BM25style rankers [41] remain widely used to fetch candidate passages. Self-RAG [17] further introduces self-reflective retrieval and critique tokens to mitigate retrieval errors. In the medical domain, Almanac [42] highlights the clinical value of retrieval augmentation over curated guideline corpora, and MedRAG [18] systematically benchmarks RAG components for medical QA. MedAgent builds on these insights but goes beyond a single-step RAG pipeline by explicitly training the agent, via GRPO, to call the evidence retrieval tool and ground every conclusion in retrieved citation identifiers.

### C. Alignment, RLHF, and GRPO

Aligning LLMs with human intent has been driven by instruction tuning [23], [24] and reinforcement learning from human feedback (RLHF) [23], [25]. A typical RLHF recipe trains a Bradley–Terry reward model [26] from pairwise preference data and optimizes the policy with PPO [43]. DPO [44] eliminates the need for an explicit reward model by directly optimizing a closed-form preference objective. Group Relative Policy Optimization (GRPO) [27], [28] is a more recent variant that replaces the value critic with a group-relative baseline computed from multiple rollouts under the same prompt, and has proven effective for reasoning-heavy tasks. We adopt GRPO as the RL backbone of MedAgent because evidence-based medical answering naturally benefits from comparing multiple candidate evidence-use trajectories. For efficient finetuning of large backbones we rely on LoRA [45], and for serving we adopt vLLM with PagedAttention [46].

### D. Medical Evaluation and Hallucination

Existing medical QA benchmarks such as MedQA [30], PubMedQA [31], and the medical subsets of MMLU [32] are predominantly multiple-choice and assume a fixed gold answer, which is ill-suited to evidence-based generation where the underlying evidence is dynamic. Med-HALT [13] and recent surveys on medical hallucination [14], [19] identify fabricated trials, statistics, and citations as the primary safety risk in medical LLMs. HealthBench [20] adopts rubric-style criteria co-designed with physicians to evaluate realistic clinical conversations. In a similar spirit, MedTrustBench employs verifiable Yes/No rubrics with positive and negative weights, together with an LLM judge calibrated against human experts [29], [47], to assess evidence faithfulness, evidence stratification, and clinical safety alongside linguistic quality.

### E. Tool-Using Agents and Reasoning

A recent line of work equips LLMs with the ability to invoke external tools and interleave reasoning with action. Chain-of-thought prompting [48] elicits intermediate reasoning steps; ReAct [21] couples reasoning traces with environment actions; and Toolformer [22] teaches LLMs to call APIs in a self-supervised manner. MedAgent can be regarded as a domain-specialized tool-using agent whose sole external tool is a four-way medical evidence retriever, and which is further trained with GRPO to align its tool usage and citation behavior with the principles of evidence-based medicine.

## III. System Overview

As illustrated in Fig. 2, MedAgent is designed as a retrieval-augmented medical agent [15], [42] rather than a standalone language model. Its training and evaluation system is organized into five layers: data, cold start, reward, reinforcement learning, and evaluation.

**Fig. 1:**
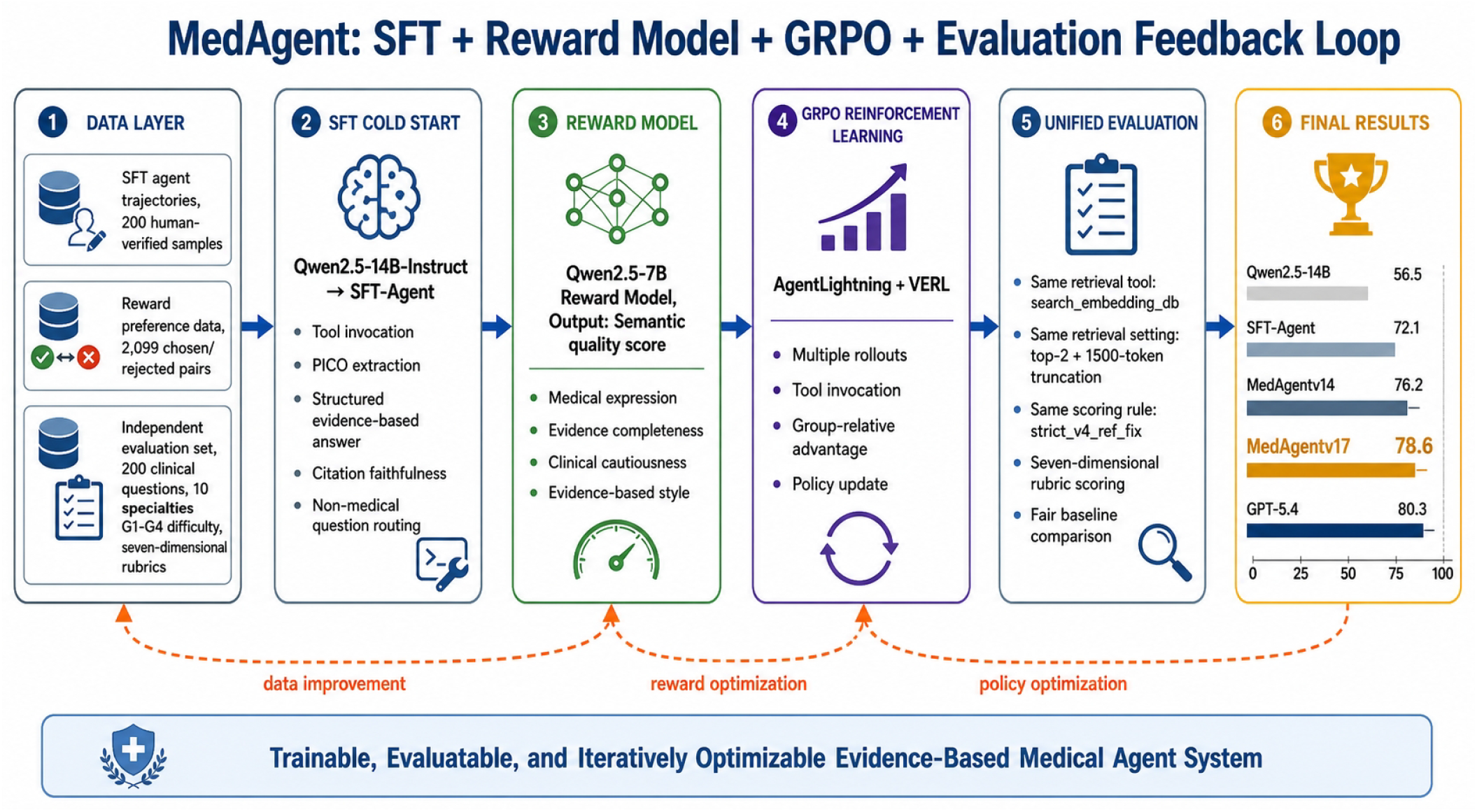
Overall pipeline of MedAgent. The system integrates SFT cold start, reward modeling, GRPO reinforcement learning, and independent rubric-based evaluation into a closed-loop evidence-based medical agent training framework.

**Fig. 2:**
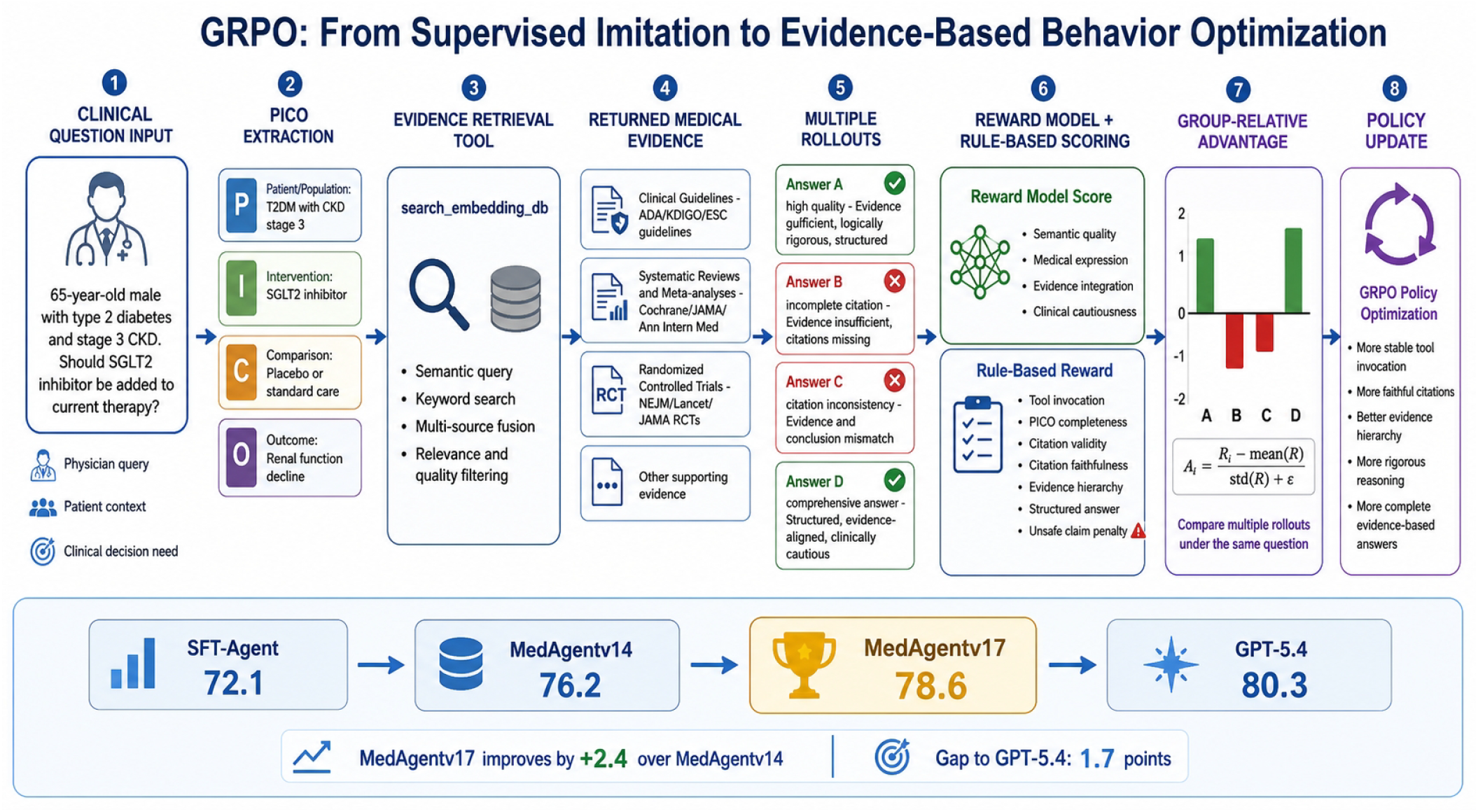
GRPO-based evidence behavior optimization. For each clinical question, the agent extracts PICO elements, invokes the evidence retrieval tool, receives multi-level medical evidence, samples multiple rollouts, and updates the policy using reward-model scores, rule-based rewards, and group-relative advantages.

**Fig. 3:**
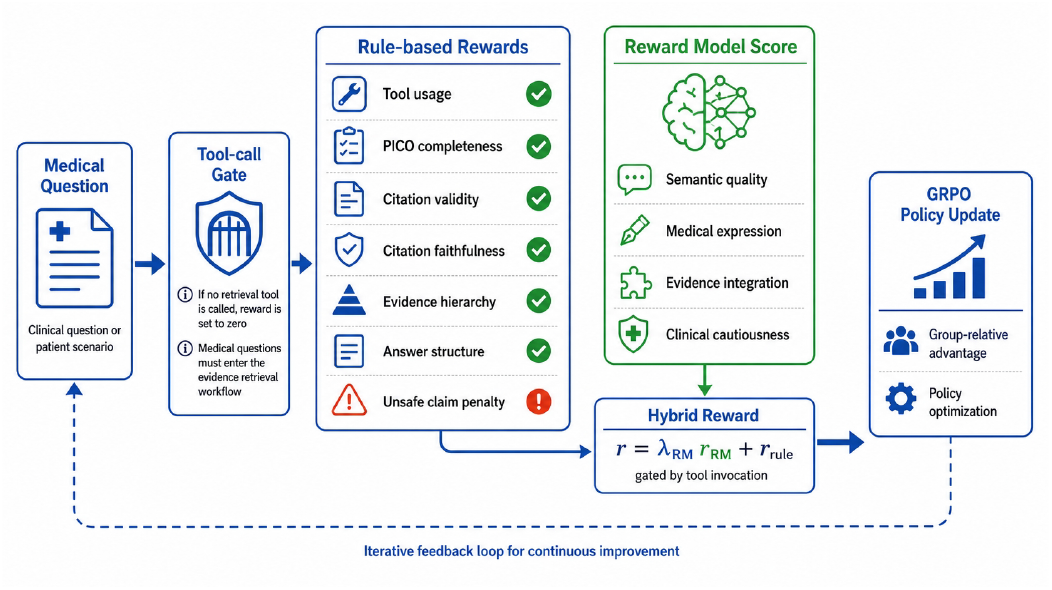
Hybrid reward design for GRPO training. Rule rewards constrain tool usage, PICO completeness, citation legality, citation faithfulness, evidence hierarchy, answer structure, and unsafe claims, while the reward model provides semantic-level quality assessment.

**Fig. 4:**
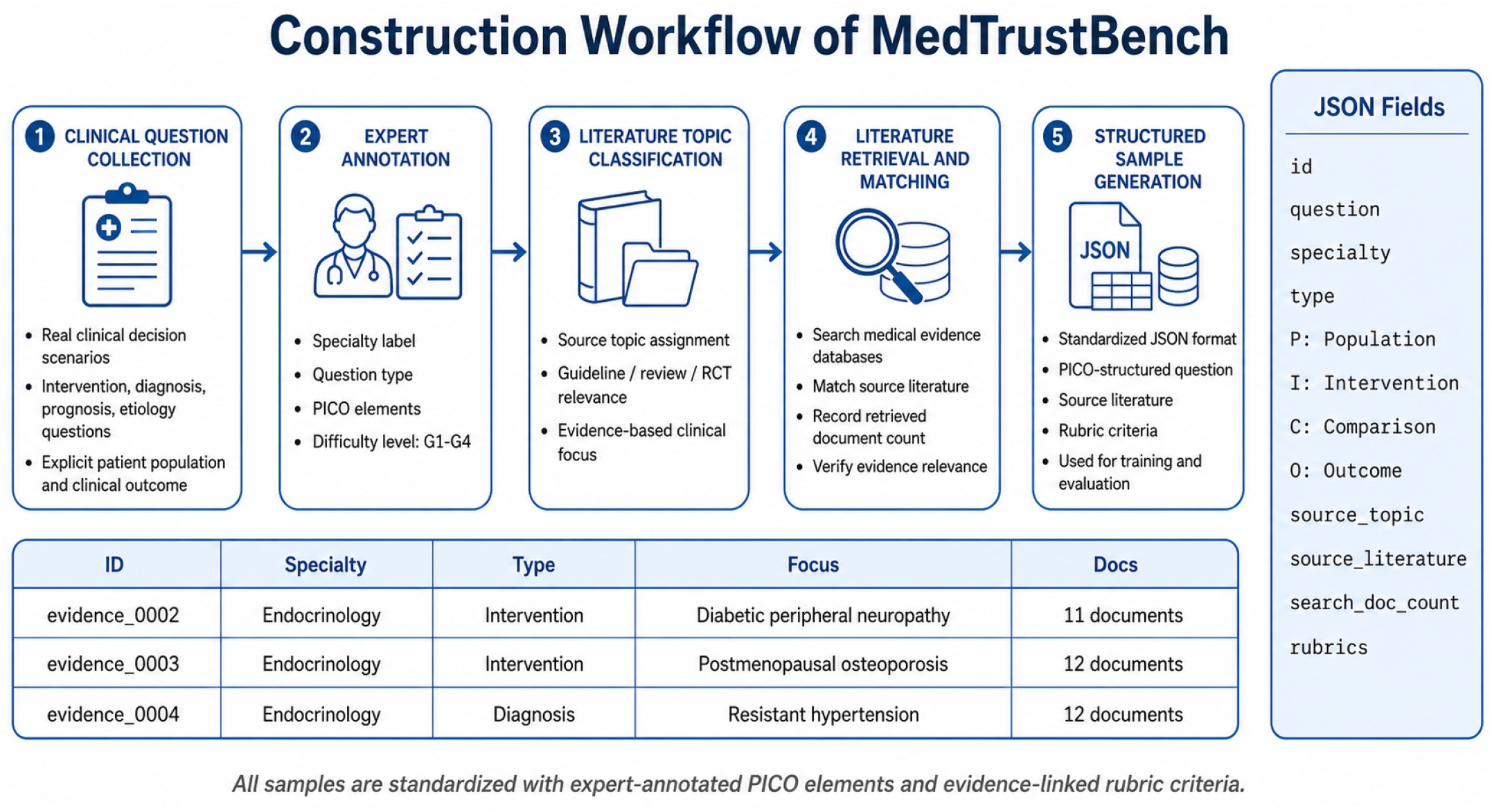
Construction workflow of MedTrustBench. Clinical questions are collected from real medical scenarios, annotated by experts with PICO elements, matched with source literature, converted into structured samples, and equipped with rubric-based scoring criteria.

**Fig. 5:**
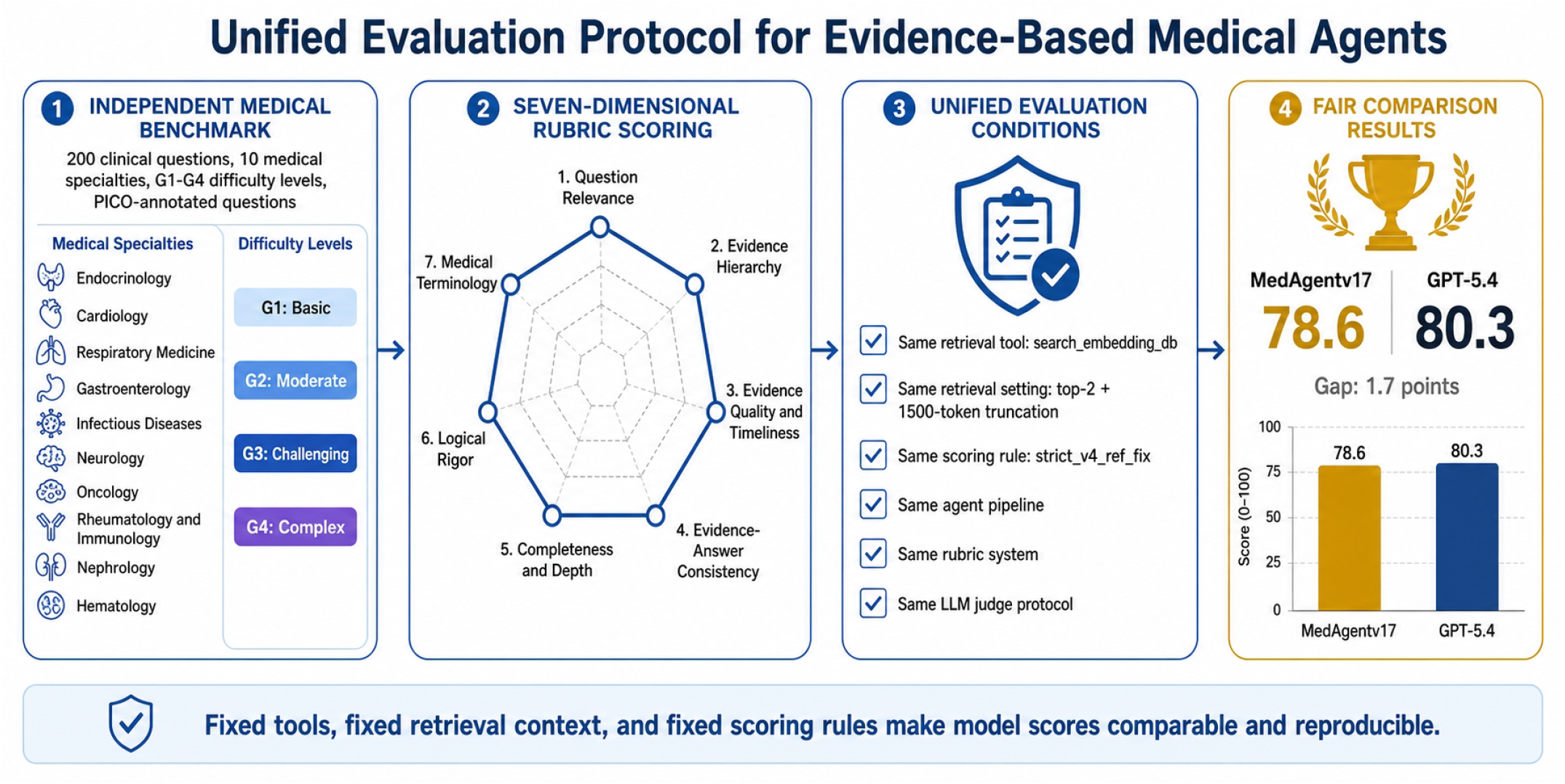
Overall model comparison. MedAgentv17 achieves 78.6 points, outperforming GPT-4.1 by 3.3 points under the same RAG pipeline and narrowing the gap to GPT-5.4 to 1.7 points.

**Fig. 6:**
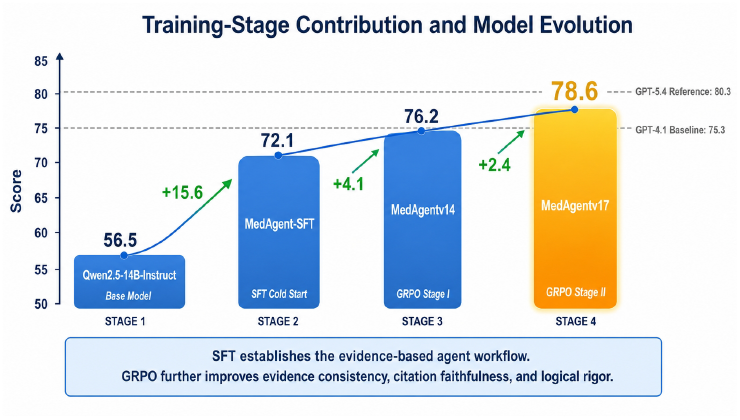
Training-stage contribution. SFT establishes the evidence-based agent workflow, while GRPO further improves evidence consistency and logical rigor.

The data layer contains three types of data. The first is the SFT cold-start dataset, consisting of 200 human-verified agent trajectories. Each trajectory captures the full interaction chain:

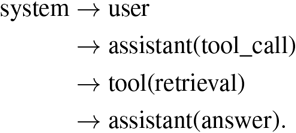

The second is the reward-model dataset, comprising 2,099 chosen/rejected preference pairs drawn from evidence-based medical answering scenarios. The third is the evaluation dataset, which contains clinical questions, PICO annotations, difficulty labels, specialty tags, source literature, and rubric criteria.

The cold-start layer applies SFT [23], [24] to transform Qwen2.5-14B-Instruct [4] from a general instruction model into an evidence-based agent. The aim is not to inject medical knowledge into the model but to instill a behavioral skeleton: when to call tools, how to emit a legal tool-call JSON, how to extract PICO elements, how to leverage retrieved evidence, how to follow a seven-part answer structure, and how to refrain from fabricated citations.

The reward layer combines a reward model with rule-based reward functions. The reward model supplies soft semantic quality scores [23], [25], whereas rule-based rewards encode explicit behaviors that can be automatically verified, such as tool usage, citation validity, citation faithfulness, evidence-hierarchy coverage, and answer-structure completeness.

The reinforcement learning layer adopts GRPO [27], [28]. In contrast to SFT, which imitates fixed reference answers, GRPO optimizes task-level reward through multiple rollouts under the same prompt. This is especially well suited to evidence-based medical agents, where alternative retrieval results, evidence combinations, and answer paths must be compared.

The evaluation layer is decoupled from the training process. It employs a fixed clinical benchmark, a fixed retrieval configuration, a fixed scoring protocol, and a seven-dimensional rubric system [20], [29], ensuring fair cross-version comparisons across model variants.

## IV. Evidence-Based Agent Formulation

Given a clinical question *q*, the agent first extracts a structured PICO representation [10]:

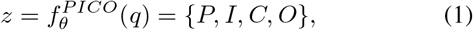

where *P, I, C*, and *O* denote Population, Intervention, Comparison, and Outcome, respectively.

The question and PICO elements are then jointly used to query the evidence retrieval tool [15], [40]:

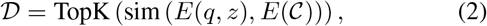

where *E*(·) denotes the embedding function, C denotes the evidence corpus, and D is the resulting set of retrieved evidence documents. In our system, the retrieval corpus is partitioned into four evidence databases: Chinese clinical guidelines, English international guidelines, systematic reviews/meta-analyses, and clinical trials [9], [11].

The final answer is then sampled from the policy model:

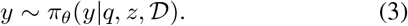

A high-quality answer should satisfy three core requirements. First, it should remain faithful to the original clinical question and its PICO structure. Second, every clinical claim should be supported by retrieved evidence. Third, each important conclusion should be traceable through valid citation identifiers returned by the retrieval tool [13], [42].

## V. SFT Cold Start

### A. Objective

The SFT cold-start stage aims to teach the model how to operate as an agent [21], [23], [24]. The base model is Qwen2.5-14B-Instruct [4], and the training data contains 200 human-verified agent trajectories. Each sample comprises a system prompt, a user clinical question, an assistant tool call, a tool-returned retrieval result, and a final assistant answer.

The system prompt specifies the model’s identity, clinical safety boundary, tool-call format, PICO extraction rule, evidence hierarchy, answer structure, and citation requirement. The assistant first emits a legal `<tool_call>` that invokes `search_embedding_db`, then produces a seven-part evidence-based answer grounded in the retrieved documents.

The SFT objective is defined over assistant tokens only:

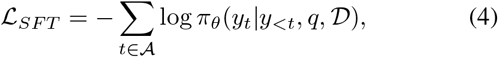

where A denotes assistant output tokens, including both tool-call JSON and final answer tokens. Tool-returned retrieval texts are excluded from the loss computation. This design discourages the model from memorizing retrieved documents and instead encourages it to learn how to invoke tools and how to organize answers using tool results.

### B. Training Configuration

All SFT experiments share the same base configuration. The base model is Qwen2.5-14B-Instruct [4], and LoRA [45] is applied with rank 32 and alpha 64. The target modules include *q proj, k proj, v proj, o proj, gate proj, up proj*, and *down proj*. The maximum sequence length is set to 6144 tokens. Training is conducted in bfloat16 precision with a warmup ratio of 0.05; packing is disabled to avoid cross-sample contamination. All experiments run on two A800 80GB GPUs.

As reported in Table I, five SFT variants are compared. V1 is under-trained and fails to produce stable tool calls. V2 forms a usable baseline, achieving stable tool invocation and structured response generation. V3, trained for five epochs at a learning rate of 1 × 10^−4^, strikes the best balance among convergence, PICO extraction quality, citation faithfulness, and answer completeness. V4 attains a lower training loss but exhibits less stable end-to-end behavior, suggesting that a higher learning rate may overfit frequent format patterns. V5 updates too conservatively and performs comparably to V2. We therefore select V3 as the SFT cold-start model for subsequent GRPO training.

**TABLE I:** SFT cold-start model selection.

| Version | Epoch | Learning Rate | Observation |
| --- | --- | --- | --- |
| V1 | 1 | $5 \times 10^{-5}$ | Under-fitted |
| V2 | 3 | $1 \times 10^{-4}$ | Stable baseline |
| V3 | 5 | $1 \times 10^{-4}$ | Best overall |
| V4 | 3 | $2 \times 10^{-4}$ | Low loss, unstable |
| V5 | 5 | $5 \times 10^{-5}$ | Conservative update |

**TABLE II:** Reward model training configuration.

| Item | Configuration |
| --- | --- |
| Backbone | Qwen2.5-7B-Instruct |
| Training samples | 2,099 pairs |
| Format | Chosen / Rejected |
| Objective | Bradley-Terry loss |
| LoRA rank / alpha | 16 / 32 |
| Dropout | 0.05 |
| Trainable parameters | ~5.04M |
| Epochs | 3 |
| Final train loss | 0.01468 |
| Deployment | FastAPI, port 8400 |

**TABLE III:** Overview of MedTrustBench.

| Item | Description |
| --- | --- |
| Dataset version | eval_dataset_final_v5.json |
| Number of questions | 200 |
| Number of specialties | 10 |
| Questions per specialty | 20 |
| Difficulty distribution | G1:18, G2:44, G3:101, G4:37 |
| Annotation format | PICO + Rubrics |
| Total rubrics | 1,187 |
| Evaluation dimensions | 7 |

**TABLE IV:** Distribution of positive and negative rubrics.

| Rubric Type | Weight | Count |
| --- | --- | --- |
| Positive | +1 | 381 |
| Positive | +2 | 477 |
| Positive | +3 | 90 |
| Negative | -2 | 67 |
| Negative | -3 | 172 |
| Total | – | 1,187 |

**TABLE V:** Seven evaluation dimensions and scoring focus.

| Dimension | Scoring Focus | Weight |
| --- | --- | --- |
| Question relevance | Alignment with clinical question and PICO | +3/ - 3 |
| Evidence hierarchy | Stratified use of guidelines, meta-analyses, RCTs | +2/ + 1 |
| Evidence quality and timeliness | Recent and authoritative evidence | +2/ + 1 |
| Evidence-answer consistency | Faithfulness between claims and evidence | -3/ + 2 |
| Completeness and depth | Benefits, risks, population, boundary | +2/ + 1 |
| Logical rigor | Evidence-analysis-conclusion chain | +2 |
| Medical terminology | Accurate and consistent medical terms | +1 |

**TABLE VI:** Rubric count by evaluation dimension.

| Dimension | Number of Rubrics |
| --- | --- |
| Question relevance | 200 |
| Evidence hierarchy | 194 |
| Evidence quality and timeliness | 194 |
| Evidence-answer consistency | 202 |
| Completeness and depth | 184 |
| Logical rigor | 113 |
| Medical terminology | 100 |
| Total | 1,187 |

**TABLE VII:** Evaluated models.

| Model | Type | Description |
| --- | --- | --- |
| Qwen2.5-14B-Instruct [4] | Base model | Original 14B instruction model without domain fine-tuning |
| MedAgent-SFT | SFT model | Qwen2.5-14B fine-tuned with evidence-based medical agent trajectories |
| MedAgentv14 | GRPO model | First-generation GRPO model based on Qwen2.5-14B, step 336 |
| MedAgentv17 | GRPO model | Second-generation GRPO model with refined rewards and citation constraints |
| GPT-4.1 [2] | Commercial baseline | OpenAI GPT-4.1 under the same RAG pipeline |
| GPT-5.4 | Commercial ceiling | OpenAI GPT-5.4 under the same RAG pipeline |

## IV. Reward Model

### A. Reward Model Dataset

The reward model is trained to judge whether an answer better satisfies evidence-based medical requirements [23], [25]. Unlike SFT, which teaches the model how to answer, the reward model learns how to evaluate answer quality.

The reward dataset comprises 2,099 pairwise preference samples in chosen/rejected format [26]. Each pair shares the same user question but contains two different assistant responses. The chosen responses generally feature a more complete PICO analysis, a clearer evidence hierarchy, more faithful citations, more cautious clinical recommendations, and stronger consistency with the underlying evidence. The rejected responses, by contrast, often suffer from missing structure, weak evidence support, unclear references, overgeneralized conclusions, or insufficient clinical reasoning.

### B. Training Objective

The reward model is built on Qwen2.5-7B-Instruct [4] and implemented as a sequence classification model with a scalar score head:

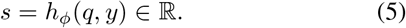

It is trained using the Bradley–Terry preference loss [23], [26]:

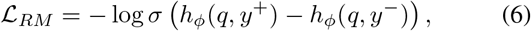

where *y*^+^ and *y*^−^ denote the chosen and rejected responses, respectively. Compared with direct preference optimization methods [44] that bypass an explicit reward model, an explicit reward model better fits our setting because it can be readily combined with verifiable rule-based rewards during GRPO.

The reward model is trained with LoRA [45] rank 16, alpha 32, dropout 0.05, and 8-bit quantization. The total number of trainable parameters is approximately 5.04 million, amounting to only about 0.07% of the 7B model. After training, the reward model is deployed as a FastAPI service that exposes `/score` and `/score-pair` endpoints for GRPO training.

## VII. GRPO Reinforcement Learning

### A. Hybrid Reward Design

Medical evidence-based generation cannot rely on a neural reward model alone. While a reward model may favor fluent and well-structured answers, fluency provides no guarantee of factual correctness or clinical safety [13], [14]. Our reward function therefore combines explicit rule-based rewards with reward-model scores.

The normalized reward-model score is given by:

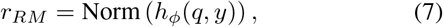

where Norm(·) maps the raw reward-model output into a bounded range.

The rule-based reward consists of several components:

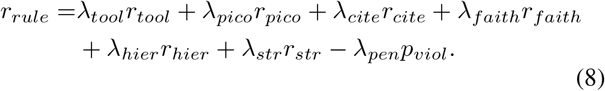

Here *r*_*tool*_ rewards successful retrieval tool usage [21], [22]; *r*_*pico*_ rewards complete PICO extraction [10]; *r*_*cite*_ checks citation format validity; *r*_*faith*_ verifies that cited identifiers originate from the retrieved documents [42]; *r*_*hier*_ rewards coverage of multiple evidence levels [9], [12]; *r*_*str*_ rewards structured answer completeness; and *p*_*viol*_ penalizes fabricated citations, unsupported references, or unsafe clinical claims [13], [19].

The final reward is gated by tool calling:

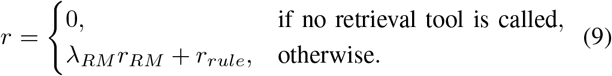

This design embodies a key clinical principle: for evidence-based medical questions, retrieval is not an optional bonus but a prerequisite. If the model answers directly from parametric memory without querying the evidence database, no positive reinforcement is granted by the evidence-based reward pipeline, regardless of linguistic quality [8], [15].

### B. GRPO Objective

For each prompt *q*, the policy samples a group of *G* rollouts [27], [28]:

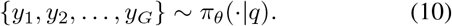

Each rollout encompasses the full agent trajectory: PICO extraction, tool call, retrieval result, and final answer. Once rewards have been obtained, the group-wise relative advantage is computed as:

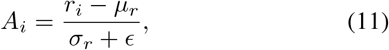

where *µ*_*r*_ and *σ*_*r*_ denote the mean and standard deviation of rewards within the same group.

The GRPO objective [27], [28], which can be viewed as a critic-free variant of PPO [43], is given by:

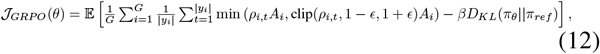

where *ρ*_*i,t*_ denotes the token-level probability ratio and *D*_*KL*_ prevents the policy from deviating excessively from the reference model [23], [43].

### C. Engineering Lines

We distinguish two GRPO training lines. The 7B engineering line builds on Qwen2.5-7B-Instruct [4] to validate the complete training, reward serving, checkpoint merging, vLLM [46] deployment, and batch evaluation pipeline. This line surfaces an important reward-design issue: if the tool-call reward is too small relative to the reward-model score, the model may achieve an apparently high training reward yet still fail to call tools reliably during pure inference.

The formal 14B line starts from the SFT cold-start model and focuses on improving evidence-answer consistency, logical rigor, citation faithfulness, and evidence hierarchy. The first-generation GRPO model is MedAgentv14 (`grpo-agent-336`), and the improved second-generation model is MedAgentv17.

## VIII MedTrustBench: Evaluation Dataset Construction

### A. Why an Independent Evaluation Dataset is Needed

Convergence of the training reward does not necessarily imply clinical reliability. A high training reward only indicates that the policy has learned to produce responses that satisfy the current reward function. It does not directly prove that the generated answer is medically accurate, faithfully grounded in evidence, or safe for clinical decision support [13], [14], [20].

In evidence-based medicine, several additional questions must be answered: Does the model cite accurate data? Does it fabricate trials or statistics? Does it correctly handle evidence conflicts? Does it stratify evidence according to the evidence hierarchy [9], [12]? Does it offer cautious recommendations when evidence is insufficient? Does it avoid over-interpreting trial results beyond the studied population? These questions cannot be fully answered by the training reward alone.

For this reason, we construct a separate evaluation benchmark that is independent of the training process. The benchmark functions as a fixed examination set for the agent and evaluates responses along multiple clinical quality dimensions rather than via a single scalar reward. The objective is to measure whether the model is genuinely reliable in evidencebased clinical reasoning, not merely whether it has learned to optimize a reward model.

### B. Design Principles

The design of MedTrustBench is inspired by recent medical AI evaluation frameworks that emphasize conditional rubric scoring and evidence-based assessment [5], [20]. Rather than comparing model answers against fixed gold answers [30]– [32], [49], our benchmark specifies what a good answer should satisfy. Each rubric is formulated as a verifiable Yes/No criterion with a fixed weight.

We adhere to three core engineering principles.

First, each rubric must be verifiable. A criterion should be judged by checking a concrete condition—for example, whether a hazard ratio matches a published trial result, whether the answer distinguishes guidelines from RCTs, or whether unsupported references are fabricated [13].

Second, the judge must be calibrated. Before any large-scale automatic evaluation, LLM-based judge results [29] should be compared with human expert judgments on a subset of samples. We use Cohen’s Kappa [47] to quantify agreement:

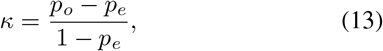

where *p*_*o*_ denotes the observed agreement and *p*_*e*_ the expected agreement by chance. The target agreement is *κ >* 0.7 before the judge is used for fully automated scoring.

Third, the evaluation set must remain frozen once finalized, ensuring fair comparison across model versions. For instance, SFT, MedAgentv14, MedAgentv17, GPT-4.1 [2], and GPT-5.4 are all evaluated on the same questions, retrieval configuration, and scoring protocol.

### C. Why Rubrics Instead of Fixed Gold Answers

Traditional question answering evaluation often hinges on gold answers and measures the similarity between model outputs and reference texts [49]. Such a methodology is problematic in evidence-based retrieval settings: the agent’s answer depends on real-time retrieved evidence, and medical literature databases are continuously updated [11]. A fixed gold answer may therefore become outdated rapidly.

In contrast, rubric-based evaluation does not prescribe exactly what the answer must say [5], [20]. It instead specifies the conditions a high-quality answer should meet. For example, a rubric may check whether the cited DAPA-CKD hazard ratio is accurate [50], whether evidence is stratified by hierarchy, or whether the answer acknowledges insufficient evidence. Because these conditions are anchored in established medical facts and evidence-based principles, they remain more stable across retrieval systems and over time.

### D. Dataset Composition

The final benchmark version, `eval_dataset_final_v5.json`, contains 200 clinical questions. It spans 10 specialties: endocrinology and metabolism, cardiovascular medicine, respiratory medicine, gastroenterology, infectious diseases, neurology, oncology, rheumatology and immunology, nephrology, and hematology. Each specialty contributes 20 questions.

Each question is annotated with standardized PICO elements [10], specialty, question type, difficulty level, source topic, source literature, and search-document count. The difficulty levels are defined as follows: G1 questions have clear and consistent guideline recommendations; G2 questions require comparison between multiple clinical options; G3 questions involve special populations, comorbidities, or benefit-risk trade-offs; and G4 questions involve conflicting evidence or inconsistent guideline recommendations.

The benchmark places strong emphasis on clinically realistic decision-making. Each question specifies a concrete patient population, a specific intervention, a comparator, and an outcome of interest. Such a design forces the agent to complete the full workflow of PICO parsing, evidence retrieval, evidence stratification, and clinical recommendation generation.

### E. Rubric Design

Each question is associated with multiple rubric criteria. Every rubric contains a criterion, a verification hint, a dimension label, and a weight. A rubric can be either positive or negative: positive rubrics reward desirable properties, while negative rubrics penalize dangerous errors such as fabricated clinical trials, unsupported statistics, or recommendations not backed by guidelines [13], [14].

The final benchmark contains 1,187 rubrics, of which 948 are positive and 239 are negative. Positive weights of +1, +2, and +3 correspond to basic requirements, important standards, and core standards, respectively. Negative weights of − 2 and − 3 correspond to severe errors and critical errors.

Negative rubrics are concentrated mainly in the evidence-answer consistency dimension. This concentration reflects the safety priority of medical AI evaluation: a fluent yet fabricated answer is more dangerous than a less polished but factually accurate one [13], [19].

### F. Seven Evaluation Dimensions

MedTrustBench assesses answers along seven dimensions. **Question relevance** measures whether the answer directly addresses the clinical question and correctly identifies PICO elements [10]. It also checks whether the answer avoids topic drift, irrelevant discussion, or misleading focus.

**Evidence hierarchy** measures whether the answer organizes evidence in accordance with evidence-based medical principles [9], [12]. It checks whether guidelines, systematic reviews, meta-analyses, and RCTs are separated and whether high-level evidence is prioritized.

**Evidence quality and timeliness** measures whether cited evidence is authoritative, relevant, and up to date. It rewards the use of current guidelines, major trials, and high-quality systematic reviews [11], [51].

**Evidence-answer consistency** measures whether answer claims are faithfully supported by the cited evidence [42]. It detects hallucinated studies, fabricated effect sizes, unsupported conclusions, and distortions of evidence [13], [14]. This is the most heavily weighted safety dimension.

**Completeness and depth** measures whether the answer covers all clinically important aspects, including benefits, risks, applicable populations, subgroups, limitations, and evidence boundaries.

**Logical rigor** measures whether the reasoning from evidence to conclusion is clear and coherent [48]. It checks whether the answer follows an evidence–analysis–conclusion chain and handles conflicting evidence in a logically sound manner.

**Medical terminology** measures whether the answer uses accurate, standardized, and consistent medical terms, including disease names, drug names, study designs, and outcome indicators.

### G. Scoring Procedure

For each model answer, the judge [29] evaluates every rubric criterion and emits a binary decision:

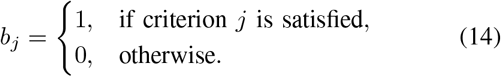

The raw score for a question is computed as:

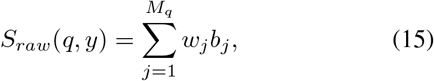

where *M*_*q*_ is the number of rubrics associated with question *q* and *w*_*j*_ is the rubric weight.

The normalized score is:

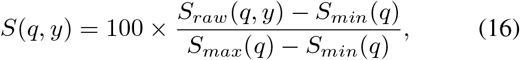

where *S*_*max*_(*q*) and *S*_*min*_(*q*) denote the maximum and minimum possible scores for that question. The final model score is the average over all questions:

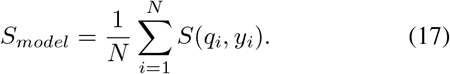

### H. Example: DAPA-CKD Question

Consider the question: “For patients with type 2 diabetes and stage 3 chronic kidney disease, can dapagliflozin, compared with placebo, delay renal function deterioration?” [50] One rubric checks question relevance: whether the answer focuses on renal outcomes of SGLT2 inhibitors in T2DM with CKD rather than drifting toward cardiovascular outcomes or unrelated drugs. A second rubric checks evidence consistency: whether the answer reports the DAPA-CKD renal composite endpoint hazard ratio as 0.61 with 95% CI 0.51–0.72 [50]. A third rubric checks whether guidelines, meta-analyses, and RCTs are presented separately [9]. A negative rubric penalizes fabricated trial names or unverifiable statistics [13].

If the model answers that DAPA-CKD demonstrated dapagliflozin’s reduction of the renal composite endpoint with HR=0.61 [50], cites CREDENCE [52] as supporting comparable SGLT2 kidney benefits, and references the KDIGO guideline recommendations [51], it earns positive scores for relevance and evidence consistency. If, however, it mixes guidelines and RCTs without stratification, it forfeits evidence-hierarchy points. As long as it fabricates no trial or statistic, no hallucination penalty applies.

This example highlights the advantage of rubric scoring: instead of asking whether the model answer is textually similar to a gold answer, the evaluation checks whether the answer satisfies concrete clinical and evidential conditions.

## IX. Unified Evaluation Protocol

All models are evaluated under the same RAG pipeline [15], [16], [18], which proceeds as:

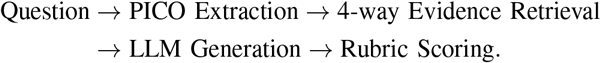

The four retrieval sources are Chinese clinical guidelines, English international guidelines, systematic reviews/meta-analyses [11], and clinical trials. Each source returns the top-2 documents, and every document is truncated to 1500 tokens. This configuration is fixed across all models.

GPT-4.1 serves as the LLM judge under a fixed rubric prompt [29]. To reduce evaluation variance, the scoring prompt explicitly instructs the judge to check each rubric independently, consult the verification hint, refrain from rewarding unsupported claims, and apply negative rubrics whenever hallucination or unsafe inference is detected [13], [14].

## X. Experiments

### A. Overall Results

Table VIII summarizes the overall comparison. The original Qwen2.5-14B-Instruct [4] reaches only 56.5 points, indicating that a general instruction model is inadequate for strict evidence-based medical reasoning, even when paired with the same retrieval pipeline. This finding confirms that retrieval alone cannot compensate for the absence of domain-specific agent alignment [18], [42].

After SFT cold start [23], MedAgent-SFT advances to 72.1 points, an improvement of 15.6 points over the base model. This is the largest single-stage gain in our pipeline, showing that high-quality agent trajectories effectively teach the model to call tools, extract PICO elements, generate structured answers, and avoid fabricated citations.

**TABLE VIII:** Overall model comparison under the same RAG pipeline.

| Rank | Model | Overall Score | Difference from GPT-4.1 |
| --- | --- | --- | --- |
| 1 | GPT-5.4 | 80.3 | +5.0 |
| 2 | MedAgentv17 | **78.6** | **+3.3** |
| 3 | MedAgentv14 | 76.2 | +0.9 |
| 4 | GPT-4.1 [2] | 75.3 | — |
| 5 | MedAgent-SFT | 72.1 | -3.2 |
| 6 | Qwen2.5-14B-Instruct [4] | 56.5 | -18.8 |

With GRPO [27], [28], MedAgentv14 climbs further to 76.2 points, surpassing GPT-4.1 [2] by 0.9 points under the same pipeline. MedAgentv17 reaches 78.6 points, outperforming GPT-4.1 by 3.3 points and narrowing the gap to GPT-5.4 to 1.7 points. The model evolution follows:

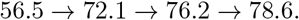

This trend demonstrates that the improvements are cumulative and remain stable across training stages.

### B. Dimension-Level Analysis

Table IX reveals that the base model already performs well on medical terminology, reaching 86.8, but lags substantially on evidence consistency, logical rigor, question relevance, and completeness. This pattern corroborates the observation that fluent medical language is not equivalent to reliable evidencebased reasoning [5], [13].

SFT substantially boosts evidence-answer consistency from 27.8 to 55.8, logical rigor from 21.7 to 49.1, and question relevance from 36.8 to 56.8. These gains indicate that the model has learned the basic evidence-based workflow. GRPO then sharpens deeper reasoning dimensions: logical rigor rises from 49.1 to 64.2, and evidence-answer consistency rises from 55.8 to 68.2. This pattern suggests that GRPO primarily strengthens evidence alignment and reasoning quality rather than merely surface formatting [27], [28].

**TABLE IX:** Dimension-level comparison among Qwen2.5-14B, MedAgent-SFT, MedAgentv14, and GPT-4.1.

| Dimension | Qwen2.5-14B | MedAgent-SFT | MedAgentv14 | GPT-4.1 |
| --- | --- | --- | --- | --- |
| Question relevance | 36.8 | 56.8 | 65.1 | 65.4 |
| Evidence hierarchy | 55.8 | 74.3 | **76.7** | 75.4 |
| Evidence quality and timeliness | 38.8 | – | 54.3 | 55.5 |
| Evidence-answer consistency | 27.8 | 55.8 | **68.2** | 66.2 |
| Completeness and depth | 31.5 | 52.8 | **56.4** | 55.0 |
| Logical rigor | 21.7 | 49.1 | 64.2 | 64.9 |
| Medical terminology | 86.8 | – | 97.4 | 99.5 |

### C. Specialty and Difficulty Analysis

MedAgentv14 performs strongly in cardiovascular medicine, rheumatology and immunology, gastroenterology, and respiratory medicine, with scores of 80.3, 80.1, 78.1, and 77.5, respectively. Neurology receives a comparatively lower score of 68.7, indicating that future training data and evaluation samples should be expanded for neurological evidence-based questions.

Across difficulty levels, MedAgentv14 attains 76.4, 76.3, 76.8, and 74.3 on G1, G2, G3, and G4 questions, respectively. This relatively balanced performance shows that the model handles not only simple guideline-consistent questions but also more complex scenarios involving special populations, benefit-risk trade-offs, and evidence uncertainty. The slightly lower G4 score is expected, as evidence-conflict questions are inherently more difficult.

### D. Training Stage Contribution

As shown in Table X, SFT yields the largest improvement because it establishes the agent’s behavioral skeleton [21], [23]. Without this stage, GRPO rollouts would contain many invalid trajectories, rendering reward signals sparse and unstable. GRPO then optimizes task-level behavior on the basis of real retrieval and multi-sample comparison [27]. MedAgentv17 achieves a further gain through refined reward functions, stronger citation-consistency constraints, and tighter alignment between training rewards and evaluation dimensions.

**TABLE X:**
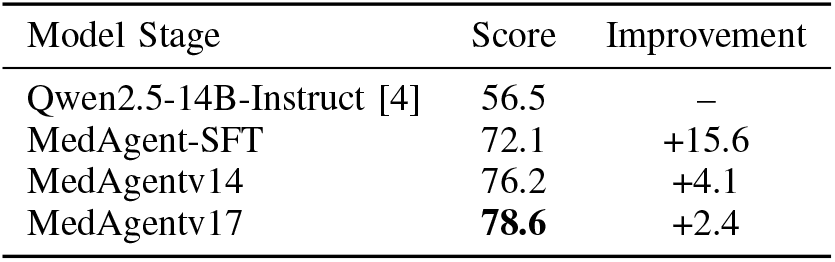
Training-stage contribution analysis.

## XI. Discussion

### A. Why Reward Convergence is Not Enough

Reward convergence is a useful signal for monitoring reinforcement learning, but it is not equivalent to clinical reliability [13], [20]. A reward model may favor long, fluent, and well-structured answers even when some claims are only weakly supported. It may also fail to detect subtle medical inaccuracies, fabricated statistics, or improper extrapolation from one patient population to another [14], [19]. An independent rubric-based evaluation is therefore essential.

The proposed evaluation benchmark decomposes answer quality into explicit clinical criteria [5], [20]. It assesses not only whether an answer is readable, but also whether it is relevant to the PICO question [10], whether the cited evidence is authoritative and recent [9], whether clinical claims are supported by the cited literature [42], whether evidence levels are properly separated [12], and whether the answer avoids hallucinated or unsafe recommendations. Such an assessment offers a far more clinically meaningful judgment than a scalar training reward.

### B. Why 14B Domain Models Can Surpass GPT-4.1

Our experimental results show that a 14B model can outperform GPT-4.1 [2] in this specialized setting. This does not imply that the 14B model is generally stronger than GPT-4.1; rather, it indicates that for a well-defined vertical task, domain-specific data [33], [34], [38], retrieval tools [15], [18], [42], reward alignment [23], [27], and strict evaluation [20] can together compensate for smaller parameter scale.

The advantage of MedAgent stems from system-level optimization. The model is not merely fine-tuned on medical text; it is trained to follow a specific evidence-based workflow [8], [10]. The retrieval tool, PICO extraction, citation constraints, reward model, rule rewards, and MedTrustBench evaluation jointly shape the final behavior.

### C. Clinical Deployment Value

A privately deployable 14B evidence-based medical agent offers several practical advantages. First, inference cost is far more controllable than long-term API usage of very large commercial models [46]. Evidence-based answers are typically long, carry multiple citations, and require long-context processing, which together drive up token-based API costs.

Second, private deployment better satisfies medical data privacy requirements. Hospitals routinely process patient records, laboratory results, medication histories, internal guidelines, and clinical pathways. Transmitting such data to external APIs may raise compliance and security concerns, whereas a local MedAgent deployment can keep all data inside the hospital network.

Third, the system is easier to customize. Hospitals can connect private knowledge bases, local clinical pathways, drug formularies, departmental guidelines [51], and expert consensus documents, and the same training and evaluation loop can subsequently drive department-specific optimization.

### D. Limitations

This work has several limitations. First, although MedTrustBench spans 10 specialties and 200 questions, rare diseases and low-frequency specialties remain underrepresented. Second, the current agent operates mainly via single-round retrieval. Real clinical reasoning may instead require iterative retrieval, evidence comparison, and conflict resolution [17], [21]. Third, although LLM-as-judge enables efficient largescale evaluation [29], expert physician calibration remains necessary. Fourth, the current citation check focuses on whether citation identifiers originate from retrieved documents. Future work should additionally verify whether each cited document semantically supports the exact claim being made [13], [42].

## XII. Future Work

Future work will proceed along five directions. First, we will expand training and evaluation data for low-performing specialties, particularly neurology and other complex clinical domains. Second, we will strengthen support for G4 evidence-conflict questions, where multiple guidelines or studies may disagree. Third, we will improve evidence quality and timeliness by refining retrieval ranking and evidence filtering [40], [41]. Fourth, we will extend the agent from single-round retrieval to multi-round retrieval and evidence comparison [17], [21]. Fifth, we will establish a hospital expert feedback loop that converts physician judgments into new SFT data, preference pairs, or GRPO reward signals [25], [44].

## XIII. Conclusion

We presented MedAgent, an evidence-based medical agent trained through SFT cold start, reward modeling, and GRPO reinforcement learning. The model is designed to execute a complete evidence-based workflow: PICO extraction, evidence retrieval, evidence stratification, citation-grounded answer generation, and quality evaluation. To rigorously evaluate the agent, we constructed MedTrustBench, an independent rubric-based benchmark featuring 200 clinical questions, 10 specialties, four difficulty levels, and 1,187 rubrics across seven dimensions.

Experimental results trace a clear model evolution path:

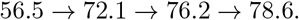

MedAgentv17 reaches 78.6 points, surpassing GPT-4.1 by 3.3 points under the same RAG pipeline and narrowing the gap to GPT-5.4 to 1.7 points. These findings validate the feasibility of the “small domain model + retrieval tool + reinforcement learning alignment” paradigm in evidence-based medicine. More importantly, MedAgent offers a deployable, auditable, privacy-preserving, and iteratively optimizable solution for hospital-oriented clinical decision support.

## Data Availability

All datasets and the trained MedAgent model produced in the present study
are publicly available online at the ModelScope repository:
https://www.modelscope.cn/profile/InfoxmedModel
The MedTrustBench evaluation benchmark, including 200 clinical questions
across 10 specialties with PICO annotations and 1,187 rubric criteria,
is openly released for reproducibility. The supervised fine-tuning
trajectories, reward model preference pairs, and evaluation scripts are
also accessible via the same repository.
No human subjects, patient records, or identifiable personal health
information were used in this study. All medical evidence referenced in
the training and evaluation pipelines was derived from publicly available
clinical guidelines, systematic reviews, meta-analyses, and randomized
controlled trials.

https://www.modelscope.cn/profile/InfoxmedModel

